# Implications of Genetic Distance to Reference and *De Novo* Genome Assembly for Clinical Genomics in Africans

**DOI:** 10.1101/2020.09.25.20201780

**Authors:** Daniel Shriner, Adebowale Adeyemo, Charles N. Rotimi

## Abstract

In clinical genomics, variant calling from short-read sequencing data typically relies on a pan-genomic, universal human reference sequence. A major limitation of this approach is that the number of reads that incorrectly map or fail to map increase as the reads diverge from the reference sequence. In the context of genome sequencing of genetically diverse Africans, we investigate the advantages and disadvantages of using a *de novo* assembly of the read data as the reference sequence in single sample calling. Conditional on sufficient read depth, the alignment-based and assembly-based approaches yielded comparable sensitivity and false discovery rates for single nucleotide variants when benchmarked against a gold standard call set. The alignment-based approach yielded coverage of an additional 270.8 Mb over which sensitivity was lower and the false discovery rate was higher. Although both approaches detected and missed clinically relevant variants, the assembly-based approach identified more such variants than the alignment-based approach. Of particular relevance to individuals of African descent, the assembly-based approach identified four heterozygous genotypes containing the sickle allele whereas the alignment-based approach identified no occurrences of the sickle allele. Variant annotation using dbSNP and gnomAD identified systematic biases in these databases due to underrepresentation of Africans. Using the counts of homozygous alternate genotypes from the alignment-based approach as a measure of genetic distance to the reference sequence GRCh38.p12, we found that the numbers of misassemblies, total variant sites, potentially novel single nucleotide variants (SNVs), and certain variant classes (*e.g*., splice acceptor variants, stop loss variants, missense variants, synonymous variants, and variants absent from gnomAD) were significantly correlated with genetic distance. In contrast, genomic coverage and other variant classes (*e.g*., ClinVar pathogenic or likely pathogenic variants, start loss variants, stop gain variants, splice donor variants, incomplete terminal codons, variants with CADD score ≥20) were not correlated with genetic distance. With improvement in coverage, the assembly-based approach can offer a viable alternative to the alignment-based approach, with the advantage that it can obviate the need to generate diverse human reference sequences or collections of alternate scaffolds.

## Introduction

The “best practice” for variant calling from short-read sequencing data relies on aligning the reads to a universal human reference (Van der Auwera et al. 2013), currently GRCh38 (https://www.ncbi.nlm.nih.gov/grc/human). There are key limitations with this alignment-based approach. One, the human reference sequence is haploid (International Human Genome Sequencing Consortium 2001). Of all possible haplotypes, the human reference sequence represents only one, and that one might not be observed in any given individual. Two, the human reference sequence was initially assembled from libraries generated from multiple individuals (International Human Genome Sequencing Consortium 2001). This assembly process yielded a mosaic, rather than consensus, haploid representation. Individual RP11, who contributed ∼70% of the sequence in GRCh38, was inferred to be an admixed African American (Schneider et al.2017). As genetic distance of a sample to the reference sequence increases, or equivalently as alignment scores decrease, reads map incorrectly or fail to map (Degner et al. 2009; Iqbal et al. 2012). This issue is expected to be worst for individuals most genetically divergent from the current human reference sequence, *e.g*., continental Africans from traditional hunter-gatherer populations in central and southern Africa as well as Oceanians (Mallick et al. 2016; Hwang et al. 2019).

Two approaches have been described to incorporate naturally occurring genetic variation into the variant calling procedure. In one approach, the current human reference sequence is customized by editing specific positions (Dewey et al. 2011; Yuan et al. 2015). In the second approach, catalogs of genetic variation are incorporated into genome graphs that improve read mapping and simplify variant calling (Paten et al. 2017; Rakocevic et al. 2019). The incorporation of alternate scaffolds in GRCh38 is an initial step in the latter approach (Schneider et al. 2017).

In large-scale genomic studies, multi-sample calling and ensemble or consensus pipeline approaches can reduce the false positive rate of variant calling, but at a cost of lower sensitivity, particularly for pathogenic variants that tend to be rare (Hwang et al. 2014; Ren et al. 2018). However, the typical clinical genomic application involves a single tested individual and not calling rare variants can result in failure to achieve a molecular diagnosis. Here, we posit that the best reference sequence for variant calling is the individual’s own genome. The use of a different reference sequence for each individual precludes multi-sample calling and yields an opportunity to reconsider pipelines of hard filtering for variant calling. Using only 100 bp paired-end read data (the typical output of current short-read sequencing technology), we formulated two pipelines for variant calling that differed only in the choice of the reference sequence: GRCh38 or the *de novo* assembly of the individual’s genome. Based on 44 continental Africans spanning a wide range of the human evolutionary tree (Mallick et al. 2016), we found that the assembly-based approach yielded coverage of 2.50 Gb, whereas the alignment-based approach yielded coverage of an additional 270.8 Mb. Furthermore, we found that the sensitivity and the false discovery rate of variant calling was identical over the fraction of the genome covered by both reference sequences. For the fraction of the genome covered by GRCh38 not covered by the *de novo* assembly, sensitivity was lower, and the false discovery rate was higher. Of particular relevance to individuals of African descent, the assembly-based approach identified four heterozygous genotypes containing the sickle allele that were missed by the alignment-based approach, reflecting a clinically important limitation of the alignment-based approach when individuals are genetically distant to the universal human reference sequence.

## Materials and Methods

### Samples, sequencing, and quality control

We retrieved 124 FASTQ files from the Sequence Read Archive for all 44 continental Africans from the Simons Genome Diversity Project (Mallick et al. 2016). The 44 continental Africans comprised two Bantu-speakers from Kenya, two Biaka from the Central African Republic, three Dinka from Sudan, two Esan from Nigeria, two Gambians, two Herero from Botswana or Namibia, four Ju/’hoansi from Namibia, two Khomani from South Africa, two Luhya from Kenya, two Luo from Kenya, two Maasai from Kenya, three Mandenka from Senegal, four Mbuti from the Democratic Republic of Congo, two Mende from Sierra Leone, two Mozabites from Algeria, two Sahrawi from Western Sahara or Morocco, one Somali from Kenya, two Tswana from Botswana or Namibia, and three Yoruba from Nigeria. These samples represent 19 ethno-linguistic groups, covering all primary language families and ranging across the entire continent. These samples also represent some of the most diverse and understudied human lineages (*e.g*., Ju/’hoansi and Khomani as well as Biaka and Mbuti) to more recently diverged groups (*e.g*., Mandenka). Five samples were sourced from saliva, five samples were sourced from blood, and the remaining 34 samples were sourced from lymphoblastoid cell lines. For 39 samples, libraries were prepared using a PCR-free protocol. For the remaining five samples, libraries were prepared using a PCR-based protocol. All samples were sequenced using 100 bp paired-end sequencing on HiSeq2000 sequencers with an average insert length of 314 bases. We performed quality control using fastp version 0.19.6 (Chen et al. 2018) and FastQC version 0.11.8 (Andrews 2010). We preprocessed the FASTQ files with fastp, using the options -y to enable the low complexity filter, -g to trim polyG tails, -x to enable polyX trimming of 3’ ends, -p to enable overrepresentation analysis, -c to enable base correction, and --length_required 100 to discard reads shorter than 100 bp.

### *De novo* assembly and quality control

We used kmergenie version 1.7048 (Chikhi and Medvedev 2014) to estimate the best *k*-mer length for *de novo* assembly of whole genomes. We then used SOAPdenovo2 release 240 (Luo et al. 2012) to perform *de novo* assembly. Since we had only one short-read library for each genome, we assembled contigs but did not assemble scaffolds. We excluded all contigs with length less than 100 bp. To evaluate the assemblies, we used QUAST version 5.0.2 (Mikheenko et al. 2018) with the following options: --large, --min-contig 100, --min-alignment 100, --extensive-mis-size 100, --fragmented, and --unaligned-part-size 1. In QUAST, a relocation is defined as a misassembly in which there is a gap between the left and right query sequences, or the left and right query sequences overlap, with the size of the gap or overlap determined by the --extensive-mis-size argument. We used the default setting of a minimum of 95% sequence identity. We also estimated gene content in the assemblies based on release 95 of the Ensembl gff3 file (ftp://ftp.ensembl.org/pub/release-95/gff3/homo_sapiens/Homo_sapiens.GRCh38.95.gff3.gz), which contains 21,492 protein-coding genes, 15,183 pseudogenes, 21,987 short non-coding RNAs, and 73 long non-coding RNAs. We aligned the assemblies to GRCh38.p12 (ftp://ftp.ncbi.nlm.nih.gov/genomes/all/GCF/000/001/405/GCF_000001405.38_GRCh38.p12/GCF_000001405.38_GRCh38.p12_genomic.fna.gz), which includes 594 sequences totaling 3.26 Gb: 25 primary chromosomal assemblies, 42 sequences localized to specific chromosomes, 126 sequences not localized to any chromosome, 140 patches, and 261 alternate scaffolds at 187 loci.

### Alignment and quality control

For each individual, we aligned the short reads using bwa mem version 0.7.17 (Li 2013) with the -M option to (1) GRCh38.p12 and (2) to the *de novo* assembly. We used samblaster version 0.1.24 (Faust and Hall 2014) to remove duplicate reads from the bam files. We used samtools version 1.9 (Li et al. 2009) with flags -f 3 and -F 3852 to filter the bam files. To reduce further false positive variant calls due to misaligned sequences, we excluded read pairs if either read contained clipped sequences, had an alignment score less than 95, or had a map quality less than 20.

### Variant calling and quality control

We used Platypus version 0.8.1 (Rimmer et al. 2014) with all default settings for variant calling and variant filters, including those for minimum read depth, allele bias, and strand bias. We used vcftools version 0.1.16 (Danecek et al. 2011) to perform quality control on the vcf files. First, we excluded variants using options --remove-filtered-all and --minQ 20. Second, we removed variants with read depth greater than 500. Lastly, we remapped variants called on alt scaffolds to the primary assembly.

### Variant annotation

We used bcftools (in samtools version 1.9) to annotate the variant calls with rsids from dbSNP version 153 (Sherry et al. 1999). We used Variant Effect Predictor (VEP) version 98 (McLaren et al. 2016) to annotate variants, with custom links to gnomAD version 3.0 (Karczewski et al. 2020), CADD version 1.5 (Rentzsch et al. 2019), and ClinVar version 20191231 (Landrum et al. 2018).

### Benchmarking

For benchmarking, we selected the Genome in a Bottle Consortium reference sample NA12878. We retrieved the integrated vcf file for NA12878 from the Genome in a Bottle Consortium ftp site (ftp://ftp-trace.ncbi.nlm.nih.gov/giab/ftp/release/NA12878_HG001/NISTv3.3.2/GRCh38/HG001_GRCh38_GIAB_highconf_CG-IllFB-IllGATKHC-Ion-10X-SOLID_CHROM1-X_v.3.3.2_highconf_PGandRTGphasetransfer.vcf.gz) (Zook et al. 2019). NA12878 is a female from the CEU population (Utah residents with ancestry from northern and western Europe). The vcf file covers 2.44 Gb of chromosomes 1 through 22 and X. We also retrieved FASTQ files containing short-read sequencing data for this individual from the Sequence Read Archive, accession SRR622457. We analyzed these read data following the steps described above, with the exception that the paired-end reads were 101 bp in length. We used asdpex version 0.3 (Jäger et al. 2016) to annotate each locus for the presence of 0, 1, or 2 alternate scaffold sequences.

## Results

### Variant Calling

The retrieved 124 FASTQ files contained 76 billion reads, of which 69 billion remained after quality control with fastp. According to FastQC quality control metrics, 29% of FASTQ files showed overrepresented sequences before quality control, whereas no file showed overrepresented sequences after quality control.

After quality control of the reads, we performed *de novo* assembly for the 44 continental African samples (Figure 1). For the five samples from blood, the mean genome size was 3.05 Gb, ranging from 3.04 to 3.06 Gb (Supplementary Table S1). Similarly, for the 34 samples from lymphoblastoid cell lines, the mean genome size was 3.06 Gb, ranging from 3.03 to 3.11 Gb (Supplementary Table S1). In contrast, for the five samples from saliva, the mean genome size was 3.27 Gb, ranging from 3.08 to 3.41 Gb (Supplementary Table S1). The average number of contigs per assembly was 10.7 million, with an average N50 of 2.7 kb (Supplementary Table S1).

**Figure 1.**
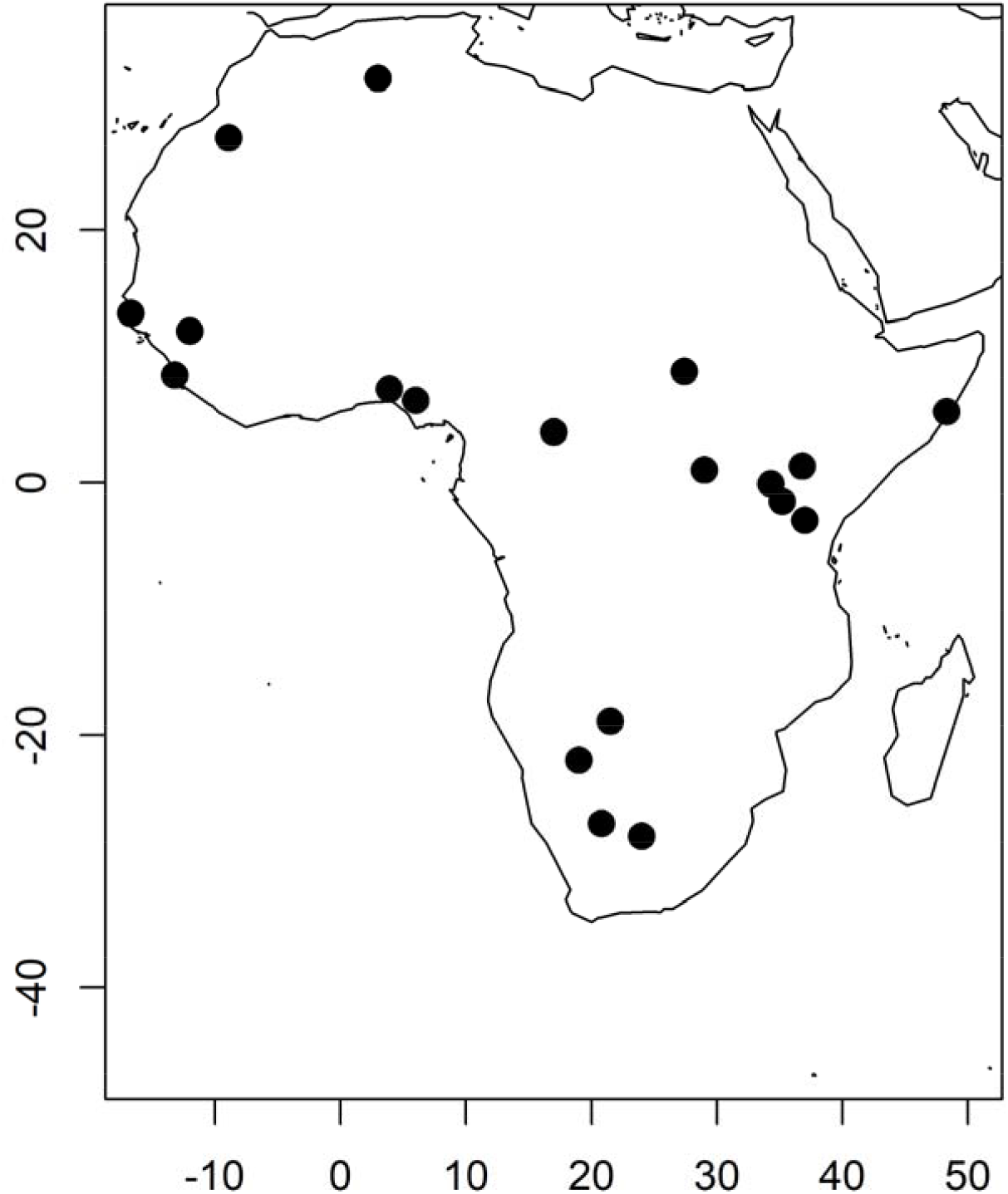
Geographical distribution of sampled individuals. The 44 continental Africans comprised two Bantu-speakers from Kenya, two Biaka from the Central African Republic, three Dinka from Sudan, two Esan from Nigeria, two Gambians, two Herero from Botswana or Namibia, four Ju/’hoansi from Namibia, two Khomani from South Africa, two Luhya from Kenya, two Luo from Kenya, two Maasai from Kenya, three Mandenka from Senegal, four Mbuti from the Democratic Republic of Congo, two Mende from Sierra Leone, two Mozabites from Algeria, two Sahrawi from Western Sahara or Morocco, one Somali from Kenya, two Tswana from Botswana or Namibia, and three Yoruba from Nigeria.

After alignment of the *de novo* assemblies to GRCh38.p12, we retained an average of 2.70 Gb, ranging from 2.66 to 2.74 Gb, yielding an average genomic coverage of 85.98%, ranging from 85.02% to 87.03% (Supplementary Table S1). On average, the *de novo* assemblies also yielded complete or partial coverage of 97.6% for protein-coding genes, 90.6% for pseudogenes, 94.9% for ncRNAs, and 98.7% for lncRNAs, indicating substantial genic coverage despite highly fractured genomes (Supplementary Table S1). The *de novo* assemblies averaged 523 kb of insertions and 752 kb of deletions relative to GRCh38.p12, with 86.6% of indels ≤5 bp (Supplementary Table S1).

We then aligned the reads to both GRCh38.p12 and the *de novo* assemblies using bwa mem. After quality control, the alignment-based approach yielded an average coverage of 2.77 Gb at an average read depth of 30.8X (Supplementary Table S2). The assembly-based approach yielded an average coverage of 2.50 Gb at an average read depth of 29.2X (Supplementary Table S2), distributed over an average of 1.58 million contigs.

Next, we performed single-sample variant calling using Platypus. The alignment-based approach yielded an average of 3.57 million variants per individual, comprising 2.19 million heterozygous variants and 1.38 homozygous variants (Supplementary Table S2). In contrast, the assembly-based approach yielded an average of 1.56 million variants per individual, of which 99.8% were heterozygous and the remainder reflected alt-aware alignment (Supplementary Table S2). The alignment-based approach yielded an average of 29,024 indels per individual, compared to 373 indels per person in the assembly-based approach (Supplementary Table S2), reflecting a severe limitation for calling indels based on highly fractured genomes.

The alignment-based approach yielded a total of 740,566 SNVs without rsids, or potentially novel variants (Figure 2), of which 90.4% were singletons. Similarly, the assembly-based approach yielded a total of 652,507 potentially novel SNVs, of which 88.1% were singletons (Figure 2). Individuals already sequenced as part of the 1000 Genomes Project (The 1000 Genomes Project Consortium 2015) yielded the fewest (∼7 to 9 thousand per individual) potentially novel SNVs and southern Africans such as Khomani yielded the most (∼60 to 80 thousand per individual) potentially novel SNVs (Supplementary Table S2).

**Figure 2.**
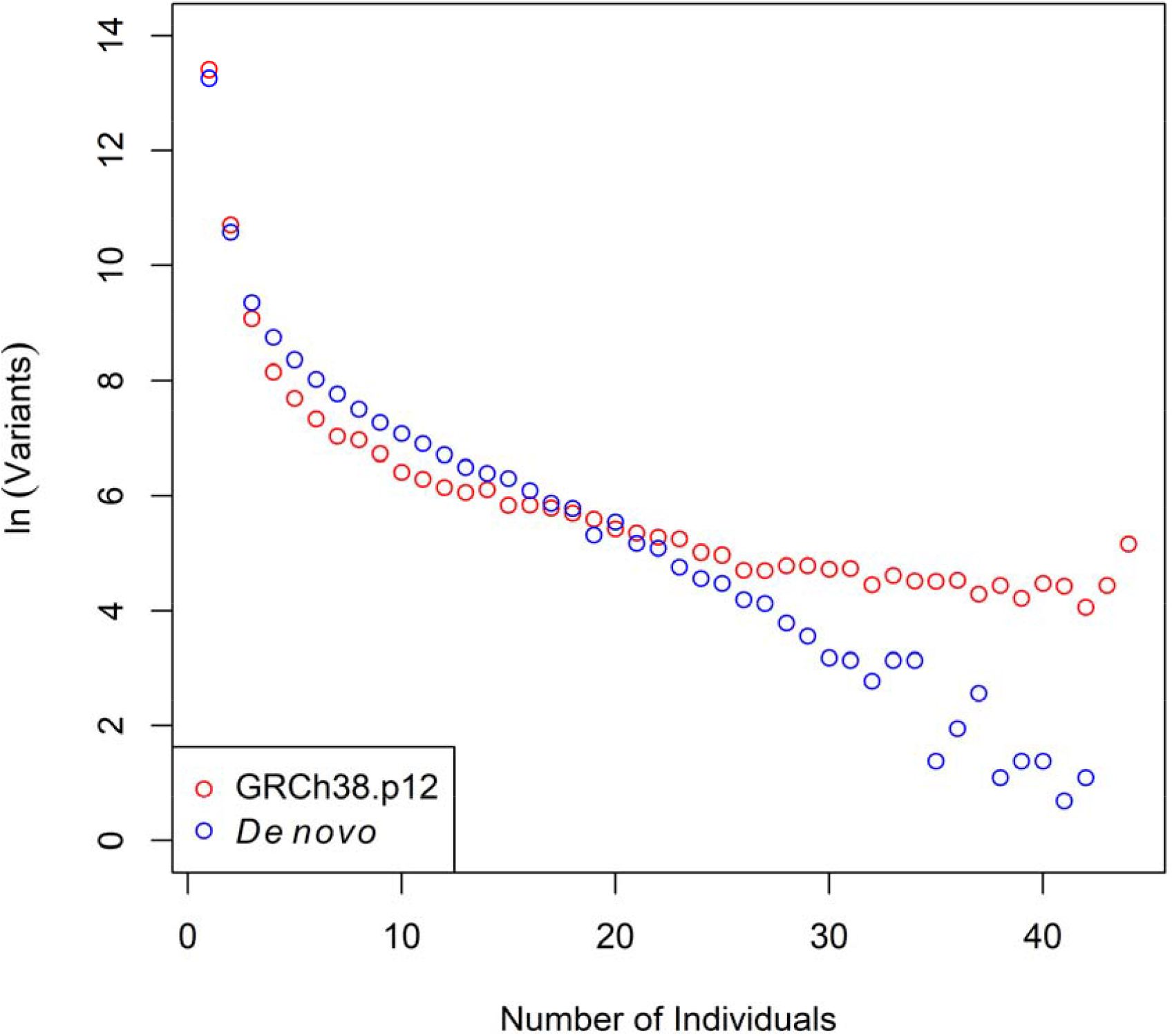
Distributions of novel variants. The *x*-axis shows the count of individuals harboring the novel variant and the *y*-axis shows a natural log-transformation of the number of variants, with novel referring to variants without rsids in dbSNP version 153.

### Variant Annotation

We annotated the variant call sets using VEP. The alignment-based approach yielded a per-individual average of 557 splice donor variants, 372 splice acceptor variants, 201 stop gains, 76 stop losses, 78 start losses, 6 incomplete terminal codons, 29,452 missense variants, and 38,509 synonymous variants (Supplementary Table S3). Annotation with ClinVar yielded an average of 12 pathogenic and 4 likely pathogenic variants per individual (Supplementary Table S3).

Annotation with CADD yielded a per-individual average of 79 variants with a phred-scaled CADD score of at least 20, reflecting the top 1% of deleteriousness (Supplementary Table S3). Annotation with gnomAD yielded a per-individual average of 38,044 African-specific variants as well as 46,433 novel variants (Supplementary Table S3). The counts for all variant types were systematically lower for the assembly-based approach when compared to the alignment approach (Supplementary Table S3). Systematic underestimation of these counts was expected because, by default, the variant call sets for the assembly-based approach did not contain homozygous genotypes.

Annotation for pathogenic or likely pathogenic ClinVar variants yielded a total of 682 heterozygous or homozygous alternate genotypes across 90 variants using the alignment-based approach and a total of 783 heterozygous or homozygous alternate genotypes across 104 variants using the assembly-based approach, after recovering homozygous alternate genotypes in the assembly-based approach (Supplementary Tables S4 and S5). Of 405 heterozygous genotypes called by the alignment-based approach, the assembly-based approach also called a heterozygous genotype for 335, yielding concordance of 82.7% (Table 2). Of 277 homozygous alternate genotypes called by the alignment-based approach, the assembly-based approach also called a homozygous alternate genotype for 253, yielding concordance of 91.3% (Table 2). Among genotypes called by both approaches, the most frequent discrepancy was a homozygous reference genotype called by the alignment-based approach *vs*. a heterozygous genotype called by the assembly-based approach (Table 2). There were 89 genotypes called by the alignment-based approach for the assembly-based approach yielded no call: 67 genotypes at 28 positions were missing due to the absence of an assembled contig, 12 genotypes at 5 positions were missing due to no reads, and 10 genotypes at 7 positions were missing due to low read depth, (Supplementary Table S5).There were 138 genotypes called by the assembly-based approach for which the alignment-based approach yielded no call: 132 genotypes at 15 positions were missing due to no reads in the cleaned bam files and 6 genotypes at 4 positions were missing due to low read depth (Supplementary Table S5).

To illustrate these findings with specific examples, we first investigated three variants of particular interest in the context of Africa. One, both approaches identified the same two individuals as heterozygous at rs2814778, the promoter-null variant of *ACKR1* that confers resistance to *Plasmodium vivax* and causes benign ethnic neutropenia. Both approaches also identified the same 32 individuals as homozygous for the alternate allele. Two, both approaches identified the same two females and one hemizygous male with one copy of the alternate allele at rs2515904, an intronic variant in *G6PD* classified as pathogenic for glucose-6-phosphate dehydrogenase deficiency. Three, the alignment-based approach did not yield any calls of the *A* allele at rs334, the causal allele for sickle cell disease, whereas the assembly-based approach yielded four heterozygotes. For these four individuals, there was selective drop-out of reads supporting the *A* allele in the alignment-based approach (Figure 3).

**Figure 3.**
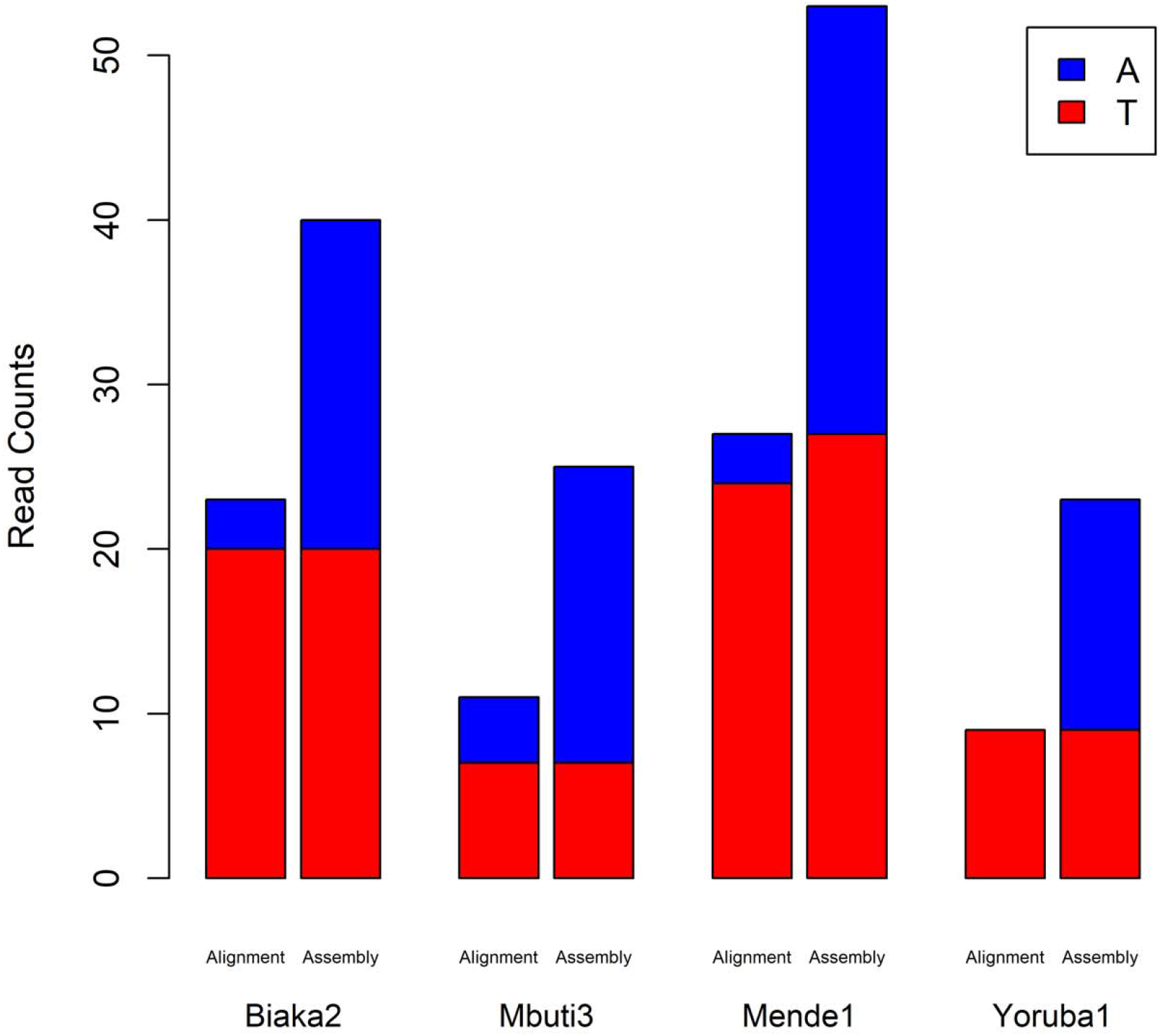
Read counts for four individuals discrepant for genotype at rs334. Read counts for the normal *T* allele are depicted in red and read counts for the sickle *A* allele are depicted in blue.

Of the pathogenic or likely pathogenic variants identified only by the alignment-based approach, we investigated two stop gain mutations, rs527236156 (R399X, detected in one heterozygote) and rs527236157 (Q407X, detected in five heterozygotes), in *KCNJ18. KCNJ18* and *KCNJ12* are paralogous genes and so detection of these two variants might be problematic due to multiple mapping of reads (Chaisson et al. 2015). In two instances, the assembly-based approach had no contig covering the position (Supplementary Table S5). In the other four instances, the assembly-based approach had a contig covering the position, but the corresponding reads were filtered out during quality control of the bam file, leaving no reads or low read depth (Supplementary Table S5). Of pathogenic or likely pathogenic variants identified only by the assembly-based approach, we investigated congenital adrenal hyperplasia, for which pathogenic variants in *CYP21A2* account for all affected individuals with 21-hydroxylase deficiency and is inherited in an autosomal recessive pattern. The assembly-based approach yielded 48 heterozygous genotypes at six different variants, whereas the alignment-based approach yielded one homozygous reference genotype and 47 missing calls, all due to no reads (Supplementary Table S5).

Last, we investigated two examples of diseases for which suspected individuals would be referred for sequencing. Alkaptonuria is an inborn error of metabolism with an autosomal recessive inheritance pattern. Both approaches identified the same individual heterozygous at rs120074172, for which the alternate allele encodes a missense variant in *HGD* classified as pathogenic. Similarly, both approaches identified the same individual heterozygous at rs730880191, for which the alternate allele encodes a missense variant in *RYR2* classified as likely pathogenic for catecholaminergic polymorphic ventricular tachycardia type 1.

### Benchmarking

We benchmarked our pipelines using the Genome in a Bottle NA12878 gold standard call set. Whereas the Genome in a Bottle Consortium integrated data from multiple platforms and sequencing technologies with a range of read lengths, our data comprised only 100 bp paired-end reads at ∼30X depth. Therefore, we expected that sensitivity would be constrained to be below 100%. By default, variant calling based on *de novo* assemblies did not yield variants homozygous for the alternate alleles. Therefore, to make fair comparisons between the two pipelines, we considered only heterozygous sites. We stratified the genome into the regions covered *vs*. not covered by the *de novo* assembly, based on the alignment of contigs to GRCh38.p12. In the regions covered by the *de novo* assembly, the alignment-based approach had higher sensitivity, 84.1% *vs*. 69.9%, whereas both pipelines had the same false discovery rate, 11.1% (Table 1). In regions not covered by the *de novo* assembly compared to regions covered by the *de novo* assembly, the alignment-based approach had 27.8% lower sensitivity and an 89.0% higher false discovery rate (Table 1). In all instances, the numbers of false negative errors exceeded the numbers of false positive errors. For variants without dbSNP rsids, both pipelines yielded similarly low sensitivities and high false discovery rates (Table 3).

**Table 1.**
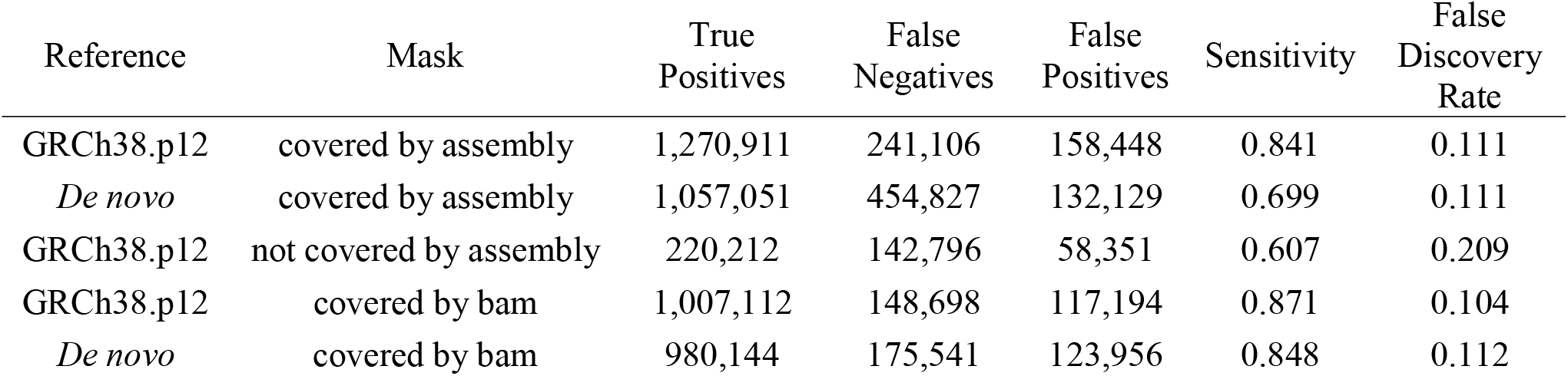
Genome-wide benchmarking of sensitivity and the false discovery rate against NA12878.

| Reference | Mask | True<br>Positives | False<br>Negatives | False<br>Positives | Sensitivity | False<br>Discovery<br>Rate |
| --- | --- | --- | --- | --- | --- | --- |
| GRCh38.p12 | covered by assembly | 1,270,911 | 241,106 | 158,448 | 0.841 | 0.111 |
| *De novo* | covered by assembly | 1,057,051 | 454,827 | 132,129 | 0.699 | 0.111 |
| GRCh38.p12 | not covered by assembly | 220,212 | 142,796 | 58,351 | 0.607 | 0.209 |
| GRCh38.p12 | covered by bam | 1,007,112 | 148,698 | 117,194 | 0.871 | 0.104 |
| *De novo* | covered by bam | 980,144 | 175,541 | 123,956 | 0.848 | 0.112 |

**Table 2.**
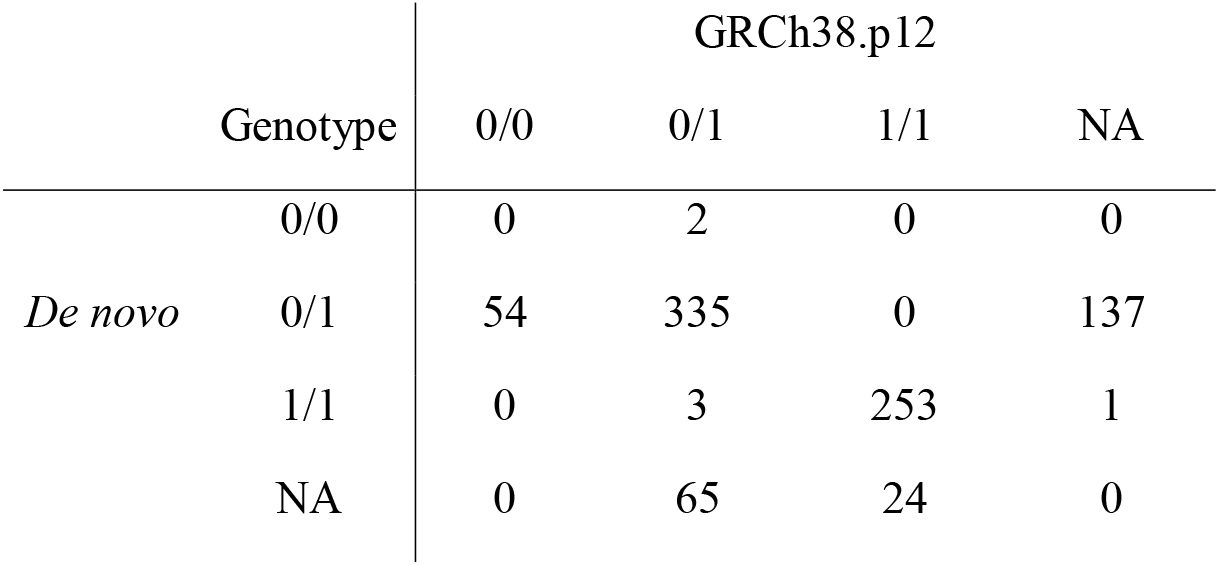
Concordance of genotypes containing at least one pathogenic or likely pathogenic ClinVar allele.

**Table 3.** Benchmarking sensitivity and the false discovery rate of novel variants against NA12878.

|  | GRCh38.p12 | *De novo* |
| --- | --- | --- |
| True Positives | 8 | 8 |
| False Negatives | 61 | 62 |
| False Positives | 104 | 3,164 |
| Sensitivity | 0.116 | 0.114 |
| False Discovery Rate | 0.929 | 0.997 |

To further investigate the reduced sensitivity of the assembly-based approach relative to the alignment-based approach for covered regions, we re-stratified the genome into regions covered *vs*. not covered by the *de novo* assembly, based on the bam file after quality control. The resulting mask comprised 1,158,921 regions covering 2,316,263,693 bp. In these regions, the alignment-based approach had a sensitivity of 87.1% and the assembly-based approach had a sensitivity of 84.8%, while the false discovery rates were 10.4% and 11.2%, respectively (Table 1). The lower sensitivity of the assembly-based approach using a mask based on aligned contigs resulted from a reduction in read depth during cleaning of the bam file, leading to some variants being filtered out due to low read depth and some variants not being called due to zero read depth.

A limitation of the benchmarking using NA12878 is that the alternate scaffolds in GRCh38.p12 were excluded during variant calling from short-read sequencing data (Zook et al. 2019). Using asdpex, we inferred that NA12878 contained 28 regions totaling 1,888 variants at which an alternate scaffold provided a better reference than the primary assembly. Of these 28 regions, 21 were heterozygous primary/alternate and 7 were homozygous alternate/alternate.

### Effect of Genetic Distance to the Universal Human Reference Sequence

We used the counts of homozygous alternate genotypes from the alignment-based approach to measure genetic distance to GRCh38.p12. We then correlated 58 statistics from the assembly, variant calling, and annotation steps to investigate the effect of genetic distance to the reference sequence. From the assembly step, only the numbers of relocations significantly correlated with genetic distance to GRCh38.p12 (Figure 4A). For protein-coding genes, pseudogenes, ncRNA genes, and lncRNA genes, the correlation with genetic distance to GRCh38.p12 for complete and partial coverage was negative and positive, respectively, but these correlations did not survive adjustment for multiple tests (Figure 4B). Similarly, genomic coverage was not significantly correlated with genetic distance to GRCh38.p12 (Figure 4B).

**Figure 4.**
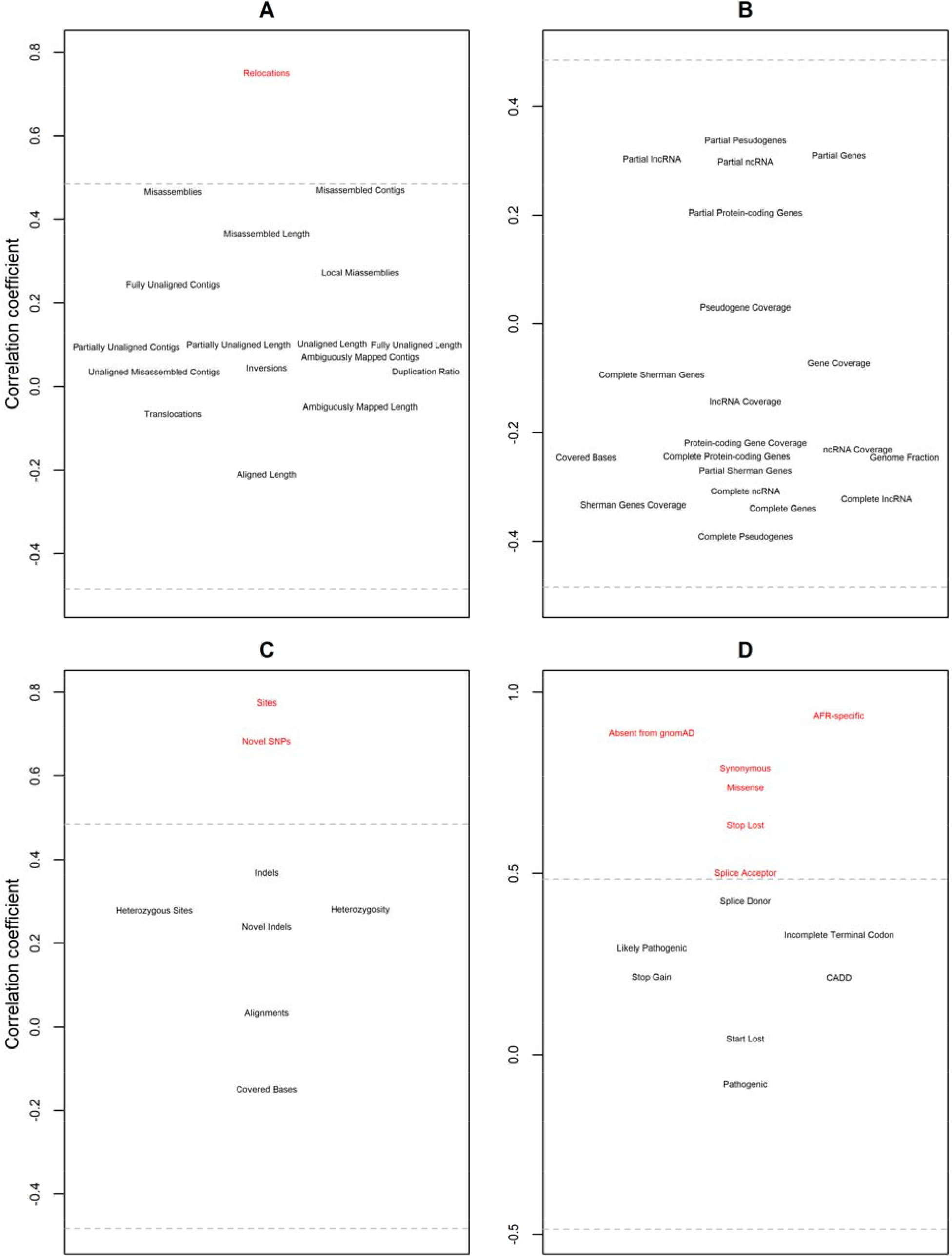
Effects of genetic distance to the universal human reference sequence. Genetic distance to GRCh38.p12 was measured using the count of variants homozygous for alternate alleles. Shown are Pearson correlation coefficients to summary statistics from (A) assembly, (B) post-assembly evaluation, (C) variant calling, and (D) variant annotation. Significance levels are indicated by the gray lines and significant correlations are depicted in red.

From the variant calling step, total (heterozygous plus homozygous) sites and potentially novel SNVs (*i.e*., without rsids in dbSNP) were both positively correlated with genetic distance to GRCh38.p12 (Figure 4C). Heterozygous sites and heterozygosity were both similar across all sub-Saharan Africans, such that neither were correlated with genetic distance to GRCh38.p12 (Figure 4C). Both pipelines indicated the heterozygosity was 16.4% lower in the North Africans compared to the sub-Saharan Africans (Supplementary Table S2).

From the VEP annotation step, the numbers of splice acceptor variants, stop codons lost, both missense and synonymous variants, variants absent from gnomAD, and variants having a non-zero allele frequency in only the gnomAD AFR group, *i.e*., being African-specific, were all positively correlated with genetic distance to GRCh38.p12 (Figure 4D). In contrast, the numbers of pathogenic or likely pathogenic variants, start codons lost, stop codons gained, splice donor variants, incomplete terminal codons, or deleterious variants (*i.e*., with a CADD score of at least 20) were not correlated with genetic distance to GRCh38.p12 (Figure 4D).

## Discussion

*De novo* assembly of ancestrally diverse humans has consistently led to the discovery of novel sequences and variants. Using 100 bp paired-end read data, we compared the alignment-based approach of variant calling with the assembly-based approach for genetically diverse individuals from across the African continent. Of note, our *de novo* assemblies are not reference quality. Despite this limitation, conditional on coverage and sufficient read depth, both approaches had comparable sensitivity and false discovery rates for SNVs. The alignment-based approach yielded higher genomic coverage but had lower sensitivity and a higher false discovery rate for regions not covered by the assembly-based approach. The assembly-based approach had very low sensitivity for indels. The assembly-based approach identified more pathogenic or likely pathogenic ClinVar variants than did the alignment-based approach.

The assembly-based approach does not involve aligning reads against a universal reference sequence. However, the assembly-based approach makes use of the current human reference sequence for two purposes. One, the current human reference sequence provides a common coordinate system. Two, the alignment of contigs from a *de novo* assembly against the current human reference sequence eliminates non-human contamination. Simultaneously, this alignment step can potentially eliminate genetically distant contigs that might harbor novel human sequences and possibly pathogenic variants. It is also possible at this alignment step that contigs could be declared misassembled and consequently eliminated. For these reasons, the assembly-based approach is sensitive to genetic distance to the universal reference sequence, with the degree of sensitivity depending on alignment scores of assembled contigs rather than short reads. Thus, like the alignment-based approach, the assembly-based approach will underperform for individuals most genetically divergent from the current human reference sequence, *e.g*., continental Africans from traditional hunter-gatherer populations in central and southern Africa.

The current human reference sequence was intended to be a pan-genome (Schneider et al. 2017), meaning that the reference contains genes that exist in at least one individual but not necessarily in all individuals. The observation that our *de novo* assemblies included a higher length of deletions than insertions is consistent with this intent. On the other hand, the assembly-based approach is biased against long indels. Previous studies have demonstrated that *de novo* assembly can identify megabases of sequence per individual missing from the human reference sequence (Li et al. 2010; Besenbacher et al. 2015; Cao et al. 2015; Seo et al. 2016; Shi et al. 2016; Maretty et al. 2017; Ameur et al. 2018). The African pan-genome from 910 individuals of African descent reportedly contains 296.5 Mb of sequence not in the current human reference, based on assembly of contigs of reads that failed to align to GRCh38.p0 (Sherman et al. 2019). These additional sequences intersected 387 known or hypothetical genes (Sherman et al. 2019), including 293 protein-coding genes, 47 non-coding RNAs, and 3 pseudogenes. Our *de novo* assemblies yielded 97.19% coverage of these genes, not just in populations that share ancestry with descendants of the Middle Passage but in populations that span the entire African continent. Given that these genes were already known, the sequences are not novel in the sense that the genes are absent from GRCh38.p12. As an alternative to the interpretation that the genomes of individuals of African descent are substantively larger than the genomes of individuals of European or other non-African descent, these sequences may have to failed to align because of high genetic distance to GRCh38.p0 and therefore may represent alternate scaffolds like those that already exist in GRCh38.

In the continental Africans we studied, we found that any individual genome differed from GRCh38.p12 by an average of 3.57 million variants. Of these variants, an average of 1.38 million reflected homozygous genotypes consisting of alternate alleles. After accounting for this fact, heterozygosity was highly similar across the sub-Saharan populations, consistent with the original description of these data (Mallick et al. 2016). Heterozygosity was 16.4% lower in North Africans, consistent with both a bottleneck during the Out of Africa migration (Henn et al. 2012) and predominantly non-sub-Saharan ancestry of North Africans (Baker et al. 2017).

Benchmarking based on the alignment of assembled contigs from NA12878 to GRCh38.p12 indicated a lower sensitivity of the assembly-based approach compared to the alignment-based approach. In contrast, benchmarking based on the cleaned bam file indicated that the sensitivity was comparable for both approaches. This result implies that some reads in the cleaned FASTQ files that were assembled into contigs were filtered out during cleaning of the bam file, leaving contigs with positions with reduced or no read depth. Thus, variants were missed either due to insufficient read depth or no coverage, respectively.

Of note, the Genome in a Bottle project did not perform alt-aware alignment and variant calling for NA12878 (Zook et al. 2019). We inferred that NA12878 had 35 haplotypes at 28 regions at which alternate scaffolds, rather than the primary assembly, should have been used as the reference sequence. Thus, it is possible that our counts of false positive variant calls are inflated and that the gold standard call set is missing variants. The benchmarking also indicated a systematic underestimation of heterozygosity, as the number of false negative errors exceeded the number of false positive errors.

At the annotation step, our results indicated a systematic bias, to a lesser extent in dbSNP and to a greater extent in gnomAD, resulting from the underrepresentation of continental Africans in catalogs of genomic variation. In the current version of gnomAD (version 3.0), the AFR label predominantly refers to African Americans (https://macarthurlab.org/2019/10/16/gnomad-v3-0/), not continental Africans, and thus gnomAD does not cover the full spectrum of genetic diversity across the African continent. Also, gnomAD used principal components analysis to assign global ancestry (https://macarthurlab.org/2019/10/16/gnomad-v3-0/), which can result in misspecified local ancestry. We caution that the estimate of the proportion of novel variants that are true from the benchmarking of an individual with European ancestry might not be applicable to individuals with diverse African ancestries and might underestimate the numbers of true novel variants. Also, all potentially novel variants remain to be validated.

The two approaches were more concordant than discordant for calling clinically relevant variants. Furthermore, the two approaches were highly concordant for homozygous alternate genotypes, which is crucial in the context of diseases with an autosomal recessive inheritance pattern. However, both approaches missed clinically relevant variants. For example, *KCNJ18* shares up to 99% nucleotide sequence identity in the coding regions with *KCNJ12* (Ryan et al. 2010). Allelic diversity in *KCNJ18* was originally mis-attributed to *KCNJ12*, until *KCNJ18* and *KCNJ12* were recognized to be paralogous genes (Ryan et al. 2010; Chaisson et al. 2015). The alignment-based approach called two stop gain variants in *KCNJ18* whereas the assembly-based approach missed both variants due to either no coverage at the assembly step or to loss of read depth during cleaning of the bam file. The fact that the variants were called in the alignment-based approach indicates that filtering out reads that mapped to multiple sites was not a problem. In contrast, the alignment-based approach identified no occurrences of the sickle allele, whereas the assembly-based approach identified four individuals heterozygous for the sickle allele. The absence of such heterozygotes in the alignment-based approach was not due to a lack of coverage but rather to selective allelic drop-out. This drop-out can be explained by the reads covering the relevant *HBB* haplotypes being too genetically distant from the current human reference sequence and therefore being filtered out based on low alignment scores. Recovering these variant calls can be achieved by lowering the threshold for alignment scores; this solution would raise sensitivity while also raising the false discovery rate.

In summary, we compared variant calling from short-read sequencing data based on the universal reference GRCh38.p12 *vs. de novo* assembly. Using simulated data, the assembly-based approach yielded lower sensitivity and a higher false discovery rate than did the alignment-based approach (Wu et al. 2017). In contrast, we found that the assembly-based approach was competitive with the alignment-based approach for calling covered SNVs. By eliminating genetic distance to a universal human reference sequence, the assembly-based approach could obviate the need to generate diverse reference sequences (https://www.genome.gov/news/news-release/NIH-funds-centers-for-advancing-sequence-of-human-genome-reference) or to keep adding alternate scaffolds to the reference sequence. The use of short-read technology (as currently prevalent) imposes an upper bound to realizing the full potential of *de novo* assembly. Despite this limitation, *de novo* assembly does provide a competitive alternative to the alignment-based approach, especially in individuals from populations that are thus far under-represented or absent from whole genome sequencing studies, with the reminder that homozygous alternate genotypes require special attention. As long read technology and diploid genome assembly become more widespread (Korlach et al. 2017; Koren et al. 2018; Miga et al. 2020; Shafin et al. 2020; Soifer et al. 2020), we anticipate that the assembly-based approach will become the standard in personalized genomic medicine.

## Data Availability

Raw data are available through the EBI European Nucleotide Archive under accession numbers PRJEB9586 and ERP010710.

https://www.ncbi.nlm.nih.gov/sra

## Acknowledgements

This work utilized the computational resources of the NIH HPC Biowulf cluster (https://hpc.nih.gov). The contents of this publication are solely the responsibility of the authors and do not necessarily represent the official view of the National Institutes of Health. This research was supported by the Intramural Research Program of the Center for Research on Genomics and Global Health (CRGGH). The CRGGH is supported by the National Human Genome Research Institute, the National Institute of Diabetes and Digestive and Kidney Diseases, the Center for Information Technology, and the Office of the Director at the National Institutes of Health (1ZIAHG200362).

## References

Ameur A, Che H, Martin M, Bunikis I, Dahlberg J, Höijer I, Häggqvist S, Vezzi F, Nordlund J, Olason P et al. 2018. De Novo Assembly of Two Swedish Genomes Reveals Missing Segments from the Human GRCh38 Reference and Improves Variant Calling of Population-Scale Sequencing Data. Genes 9: 486.

Andrews S. 2010. FastQC: a quality control tool for high throughput sequence data. Version 0.11.8. Retrieved from https://www.bioinformatics.babraham.ac.uk/projects/fastqc.

Baker JL, Rotimi CN, Shriner D. 2017. Human ancestry correlates with language and reveals that race is not an objective genomic classifier. Sci Rep 7: 1572.

Besenbacher S, Liu S, Izarzugaza JM, Grove J, Belling K, Bork-Jensen J, Huang S, Als TD, Li S, Yadav R et al. 2015. Novel variation and de novo mutation rates in population-wide de novo assembled Danish trios. Nat Commun 6: 5969.

Cao H, Wu H, Luo R, Huang S, Sun Y, Tong X, Xie Y, Liu B, Yang H, Zheng H et al. 2015. De novo assembly of a haplotype-resolved human genome. Nat Biotechnol 33: 617–622.

Chaisson MJP, Wilson RK, Eichler EE. 2015. Genetic variation and the de novo assembly of human genomes. Nat Rev Genet 16: 627–640.

Chen S, Zhou Y, Chen Y, Gu J. 2018. fastp: an ultra-fast all-in-one FASTQ preprocessor. Bioinformatics 34: i884–i890.

Chikhi R, Medvedev P. 2014. Informed and automated k-mer size selection for genome assembly. Bioinformatics 30: 31–37.

The 1000 Genomes Project Consortium. 2015. A global reference for human genetic variation. Nature 526: 68–74.

Danecek P, Auton A, Abecasis G, Albers CA, Banks E, DePristo MA, Handsaker RE, Lunter G, Marth GT, Sherry ST et al. 2011. The variant call format and VCFtools. Bioinformatics 27: 2156–2158.

Degner JF, Marioni JC, Pai AA, Pickrell JK, Nkadori E, Gilad Y, Pritchard JK. 2009. Effect of read-mapping biases on detecting allele-specific expression from RNA-sequencing data. Bioinformatics 25: 3207–3212.

Dewey FE, Chen R, Cordero SP, Ormond KE, Caleshu C, Karczewski KJ, Whirl-Carrillo M, Wheeler MT, Dudley JT, Byrnes JK et al. 2011. Phased whole-genome genetic risk in a family quartet using a major allele reference sequence. PLOS Genet 7: e1002280.

Faust GG, Hall IM. 2014. SAMBLASTER: fast duplicate marking and structural variant read extraction. Bioinformatics 30: 2503–2505.

Henn BM, Cavalli-Sforza LL, Feldman MW. 2012. The great human expansion. Proc Natl Acad Sci U S A 109: 17758–17764.

Hwang K-B, Lee I-H, Li H, Won D-G, Hernandez-Ferrer C, Negron JA, Kong SW. 2019. Comparative analysis of whole-genome sequencing pipelines to minimize false negative findings. Sci Rep 9: 3219.

Hwang K-B, Lee I-H, Park J-H, Hambuch T, Choe Y, Kim M, Lee K, Song T, Neu MB, Gupta N et al. 2014. Reducing false-positive incidental findings with ensemble genotyping and logistic regression based variant filtering methods. Hum Mutat 35: 936–944.

International Human Genome Sequencing Consortium. 2001. Initial sequencing and analysis of the human genome. Nature 409: 860–921.

Iqbal Z, Caccamo M, Turner I, Flicek P, McVean G. 2012. De novo assembly and genotyping of variants using colored de Bruijn graphs. Nat Genet 44: 226–232.

Jäger M, Schubach M, Zemojtel T, Reinert K, Church DM, Robinson PN. 2016. Alternate-locus aware variant calling in whole genome sequencing. Genome Med 8: 130.

Karczewski KJ, Francioli LC, Tiao G, Cummings BB, Alföldi J, Wang Q, Collins RL, Laricchia KM, Ganna A, Birnbaum DP et al. 2020. The mutational constraint spectrum quantified from variation in 141,456 humans. Nature 581: 434–443.

Koren S, Rhie A, Walenz BP, Dilthey AT, Bickhart DM, Kingan SB, Hiendleder S, Williams JL, Smith TPL, Phillippy AM. 2018. De novo assembly of haplotype-resolved genomes with trio binning. Nat Biotechnol 36: 1174–1182.

Korlach J, Gedman G, Kingan SB, Chin C-S, Howard JT, Audet J-N, Cantin L, Jarvis ED. 2017. De novo PacBio long-read and phased avian genome assemblies correct and add to reference genes generated with intermediate and short reads. Gigascience 6: 1–16.

Landrum MJ, Lee JM, Benson M, Brown GR, Chao C, Chitipiralla S, Gu B, Hart J, Hoffman D, Jang W et al. 2018. ClinVar: improving access to variant interpretations and supporting evidence. Nucleic Acids Res 46: D1062–D1067.

Li H. 2013. Aligning sequence reads, clone sequences and assembly contigs with BWA-MEM. arXiv 1303.3997v2 [q-bio.GN].

Li H, Handsaker B, Wysoker A, Fennell T, Ruan J, Homer N, Marth G, Abecasis G, Durbin R, GPDP Subgroup. 2009. The Sequence Alignment/Map format and SAMtools. Bioinformatics 25: 2078–2079.

Li R, Li Y, Zheng H, Luo R, Zhu H, Li Q, Qian W, Ren Y, Tian G, Li J et al. 2010. Building the sequence map of the human pan-genome. Nat Biotechnol 28: 57–63.

Luo R, Liu B, Xie Y, Li Z, Huang W, Yuan J, He G, Chen Y, Pan Q, Liu Y et al. 2012. SOAPdenovo2: an empirically improved memory-efficient short-read de novo assembler. Gigascience 1: 18.

Mallick S, Li H, Lipson M, Mathieson I, Gymrek M, Racimo F, Zhao M, Chennagiri N, Nordenfelt S, Tandon A et al. 2016. The Simons Genome Diversity Project: 300 genomes from 142 diverse populations. Nature 538: 201–206.

Maretty L, Jensen JM, Petersen B, Sibbesen JA, Liu S, Villesen P, Skov L, Belling K, Theil Have C, Izarzugaza JMG et al. 2017. Sequencing and de novo assembly of 150 genomes from Denmark as a population reference. Nature 548: 87–91.

McLaren W, Gil L, Hunt SE, Riat HS, Ritchie GRS, Thormann A, Flicek P, Cunningham F. 2016. The Ensembl Variant Effect Predictor. Genome Biol 17: 122.

Miga KH, Koren S, Rhie A, Vollger MR, Gershman A, Bzikadze A, Brooks S, Howe E, Porubsky D, Logsdon GA et al. 2020. Telomere-to-telomere assembly of a complete human X chromosome. Nature 585: 79–84.

Mikheenko A, Prjibelski A, Saveliev V, Antipov D, Gurevich A. 2018. Versatile genome assembly evaluation with QUAST-LG. Bioinformatics 34: i142–i150.

Paten B, Novak AM, Eizenga JM, Garrison E. 2017. Genome graphs and the evolution of genome inference. Genome Res 27: 665–676.

Rakocevic G, Semenyuk V, Lee W-P, Spencer J, Browning J, Johnson IJ, Arsenijevic V, Nadj J, Ghose K, Suciu MC et al. 2019. Fast and accurate genomic analyses using genome graphs. Nat Genet 51: 354–362.

Ren Y, Reddy JS, Pottier C, Sarangi V, Tian S, Sinnwell JP, McDonnell SK, Biernacka JM, Carrasquillo MM, Ross OA et al. 2018. Identification of missing variants by combining multiple analytic pipelines. BMC Bioinformatics 19: 139.

Rentzsch P, Witten D, Cooper GM, Shendure J, Kircher M. 2019. CADD: predicting the deleteriousness of variants throughout the human genome. Nucleic Acids Res 47: D886–D894.

Rimmer A, Phan H, Mathieson I, Iqbal Z, Twigg SRF, WGS500 Consortium, Wilkie AOM, McVean G, Lunter G. 2014. Integrating mapping-, assembly- and haplotype-based approaches for calling variants in clinical sequencing applications. Nat Genet 46: 912–918.

Ryan DP, MRD da Silva, Soong TW, Fontaine B, Donaldson MR, Kung AWC, Jongjaroenprasert W, Liang MC, Khoo DHC, Cheah JS et al. 2010. Mutations in potassium channel Kir2.6 cause susceptibility to thyrotoxic hypokalemic periodic paralysis. Cell 140: 88–98.

Schneider VA, Graves-Lindsay T, Howe K, Bouk N, Chen H-C, Kitts PA, Murphy TD, Pruitt KD, Thibaud-Nissen F, Albracht D et al. 2017. Evaluation of GRCh38 and de novo haploid genome assemblies demonstrates the enduring quality of the reference assembly. Genome Res 27: 849–864.

Seo J-S, Rhie A, Kim J, Lee S, Sohn M-H, Kim C-U, Hastie A, Cao H, Yun J-Y, Kim J et al. 2016. De novo assembly and phasing of a Korean human genome. Nature 538: 243–247.

Shafin K, Pesout T, Lorig-Roach R, Haukness M, Olsen HE, Bosworth C, Armstrong J, Tigyi K, Maurer N, Koren S et al. 2020. Nanopore sequencing and the Shasta toolkit enable efficient de novo assembly of eleven human genomes. Nat Biotechnol 38: 1044–1053.

Sherman RM, Forman J, Antonescu V, Puiu D, Daya M, Rafaels N, Boorgula MP, Chavan S, Vergara C, Ortega VE et al. 2019. Assembly of a pan-genome from deep sequencing of 910 humans of African descent. Nat Genet 51: 30–35.

Sherry ST, Ward M, Sirotkin K. 1999. dbSNP-database for single nucleotide polymorphisms and other classes of minor genetic variation. Genome Res 9: 677–679.

Shi L, Guo Y, Dong C, Huddleston J, Yang H, Han X, Fu A, Li Q, Li N, Gong S et al. 2016. Long-read sequencing and de novo assembly of a Chinese genome. Nat Commun 7: 12065.

Soifer I, Fong NL, Yi N, Ireland AT, Lam I, Sooknah M, Paw JS, Peluso P, Concepcion GT, Rank D et al. 2020. Fully phased sequence of a diploid human genome determined de novo from the DNA of a single individual. G3: Genes|Genomes|Genetics in press.

Van der Auwera GA, Carneiro MO, Hartl C, Poplin R, Del Angel G, Levy-Moonshine A, Jordan T, Shakir K, Roazen D, Thibault J et al. 2013. From FastQ data to high confidence variant calls: the Genome Analysis Toolkit best practices pipeline. Curr Protoc Bioinformatics 43: 11.10.11-11.10.33.

Wu L, Yavas G, Hong H, Tong W, Xiao W. 2017. Direct comparison of performance of single nucleotide variant calling in human genome with alignment-based and assembly-based approaches. Sci Rep 7: 10963.

Yuan S, Johnston HR, Zhang G, Li Y, Hu Y-J, Qin ZS. 2015. One Size Doesn’t Fit All - RefEditor: Building Personalized Diploid Reference Genome to Improve Read Mapping and Genotype Calling in Next Generation Sequencing Studies. PLOS Comput Biol 11: e1004448.

Zook JM, McDaniel J, Olson ND, Wagner J, Parikh H, Heaton H, Irvine SA, Trigg L, Truty R, McLean CY et al. 2019. An open resource for accurately benchmarking small variant and reference calls. Nat Biotechnol 37: 561–566.

